# Development and validation of a high throughput *Neisseria gonorrhoeae* genotyping method

**DOI:** 10.1101/2023.01.09.23284302

**Authors:** Kohji Komori, Kotaro Aoki, Yoshikazu Ishii, Ken Shimuta, Makoto Ohnishi, Kazuhiro Tateda

## Abstract

**Background:** *Neisseria gonorrhoeae* genotyping by whole-genome sequencing (WGS) is expensive for a large sample set, a less expensive and more efficient genotyping method is required. We developed a high-throughput genotyping method for *N. gonorrhoeae* to improve molecular epidemiological typing and antimicrobial-resistant identification in *N. gonorrhoeae* antimicrobial susceptibility surveillance.

**Methods:** We used multiplex-tailed PCR to amplify and sequence 15 alleles from multilocus sequence typing (MLST), *N. gonorrhoeae* multiantigen sequence typing (NG-MAST), and *N. gonorrhoeae* sequence typing for antimicrobial resistance (NG-STAR). After indexing-PCR, we sequenced the DNA library using the MiSeq platform (Illumina). Sequencing reads were *de novo* assembly or constructing consensus sequences of alleles, then assigned sequence type. We used 54 previously characterized strains of *N. gonorrhoeae* and WGS data to validate our method.

**Results:** The allele identification results of MLST and NG-STAR in all strains agreed with the draft WGS. However, in NG-MAST, only 35 strains agreed. Disagreement was found in the NG-MAST of *porB* in 15 strains and of *tbpB* in seven strains. QRDR analysis perfectly predicted levofloxacin resistance. But was less successful in predicting reduced susceptibility or resistance phenotype to penicillin G, cefixime, or ceftriaxone using *penA, porB, ponA*, or *mtrR* alleles.

**Conclusions:** The successful performance in MLST and NG-STAR of our method was validated in this study. This method may be useful for large-scale genotyping for *N. gonorrhoeae* surveillance in a cost- and labor-saving manner. Phenotypic prediction of antimicrobial susceptibility by combining multiple alleles may be necessary for other than fluoroquinolones.

## Introduction

The World Health Organization (WHO) estimated that there were 87 million new global cases of gonorrhea in 2016^1^. Furthermore, the increasing frequency of antimicrobial resistant (AMR) *Neisseria gonorrhoeae*, the bacterium that causes gonorrhea, is a serious public health concern^2,3^. Historically, resistance to penicillin G, fluoroquinolones, oral cephalosporins, ceftriaxone, and azithromycin has been problematic in the treatment of gonorrhea^4^. Ceftriaxone, an extended-spectrum cephalosporin or combination with azithromycin, are used as a first-line option for treating gonorrhea^5,6^. However, in low- or middle-income countries, empiric therapy with inappropriate spectrum or dose oral antimicrobial agents such as cefixime, levofloxacin, or azithromycin that are over-the-counter systems for symptom-based sexually transmitted infections may select for ceftriaxone- or azithromycin-resistant *N. gonorrhoeae*^4,7–9^. Understanding the development and spreading manor of AMR *N. gonorrhoeae* and antimicrobial resistant mechanisms are important to guide the appropriate use of the treatment option of gonorrhea and to motivate the development of novel agents to circumvent resistance.

One of the predominant mechanisms leading to resistance to ceftriaxone is a reduced affinity of penicillin binding protein 2 that causes mosaic-like mutations to cephalosporins^10,11^. In 2009, a high-level ceftriaxone-resistant *N. gonorrhoeae* strain reported in Japan called H041 and WHO X severely limited the ability to treat gonorrhea^12,13^. The first failure with combination therapy with both ceftriaxone and azithromycin was reported in the United Kingdom in 2016^14^. Therefore, the clonal spread of high-level ceftriaxone resistance alone and combinational resistance to both ceftriaxone and azithromycin should be carefully monitored.

Whole-genome sequencing (WGS) using a massive parallel sequencer (MPS) can simultaneously analyze multiple typing methods^15^, such as multilocus sequence typing (MLST)^16,17^, *N. gonorrhoeae* multi-antigen sequence typing (NG-MAST)^18^, and *N. gonorrhoeae* sequence typing for antimicrobial resistance (NG-STAR) scheme for 7 alleles associated with cephalosporins, fluoroquinolones, penicillins, and macrolides^19^. WGS using MPS is widely used and allows for a comprehensive analysis, but its cost per strain is too expensive for large-scale surveillance^15,18,20,21^. Hence, a less expensive and more efficient AMR *N. gonorrhoeae* genotyping method is required.

To address this need, we developed and validated a high throughput method that takes full advantage of the features of the MiSeq (Illumina, San Diego, CA, USA) sequence length that is commonly used in the analysis of bacteria.

## Materials and Methods

### Primers and condition for multiplex-tailed PCR

The sequences of the two primer sets and the concentrations for multiplex-tailed PCR that amplifies 15 alleles of MLST, NG-MAST, and NG-STAR are shown in Table S1. The primer design was a consensus sequence referring the genome sequence data of the 14 WHO *N. gonorrhoeae* reference strains^21^. We prepared primers with a tail sequence added to the 5’ end based on the 16S rRNA amplicon sequence preparation protocol from Illumina^22^. If the target region was longer than the MiSeq merged read (approximately 450 bp), it was amplified with two pairs of primers (left and right side) per region (Figure 1). Template DNA was extracted by boiling method that from the bacterial cell suspension in sterilized water by heating at 95°C for 15 minutes and then centrifuged. Primer concentrations were adjusted to ensure equal amplification efficiency for each allele, and validation was performed by quantitative PCR with KAPA SYBR FAST qPCR Master Mix (2×) (Nippon Genetics Co., Ltd., Tokyo, Japan). We used TaKaRa Ex Premier DNA Polymerase (TaKaRa Bio, Shiga, Japan) for multiplex-tailed PCR with an initial incubation at 94°C for 5 min, followed by 30 cycles at 98°C for 10 s, 50°C for 15 s, 68°C for 30 s, and then 68°C for 1 min. The left and right multiplex PCR products were then mixed and purified using Agencourt AMPure XP (Beckman Coulter, Inc., CA, USA).

**Figure 1:**
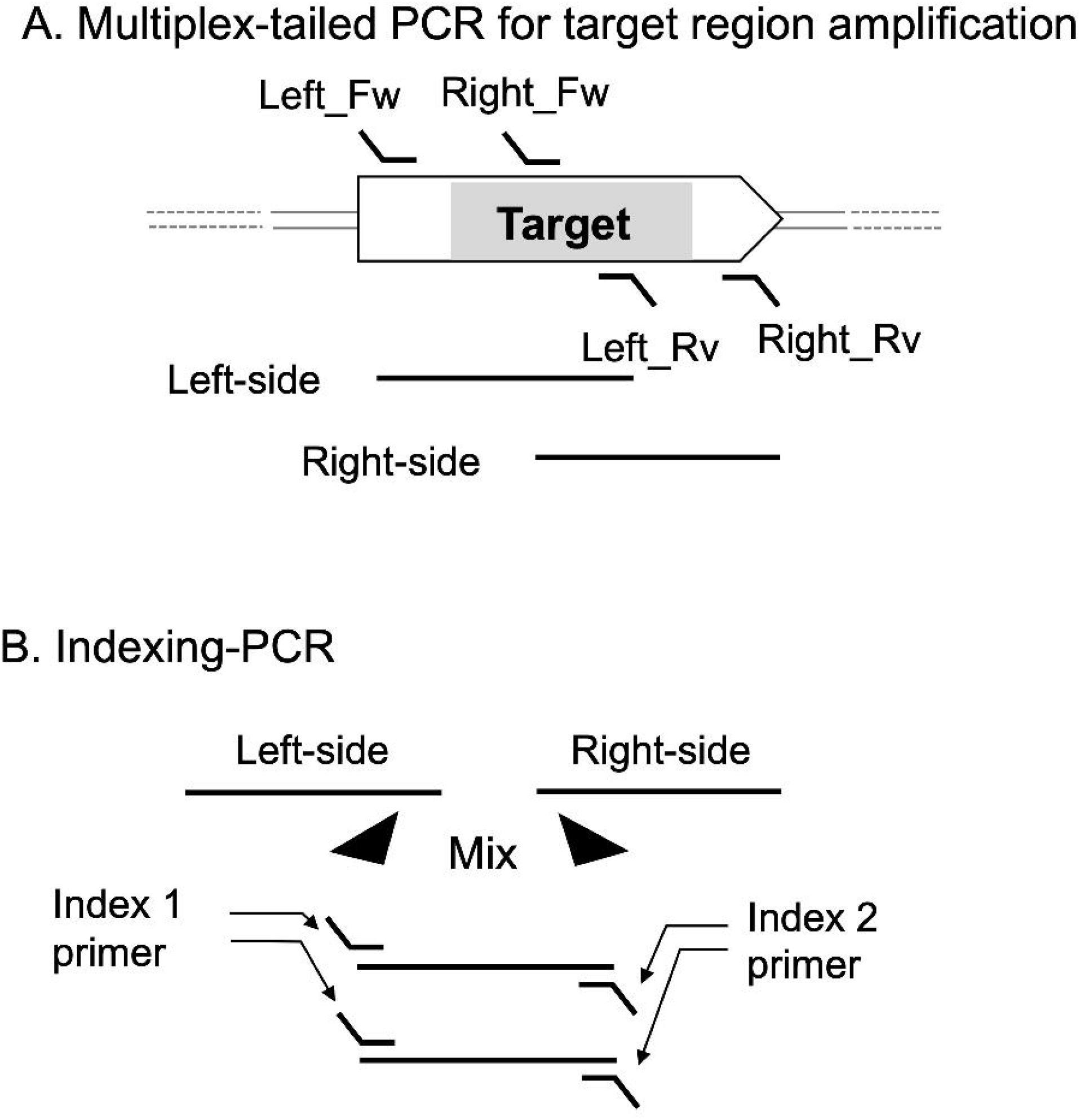
Model of MiSeq sequencing library preparation for MLST, NG-MAST, and NG-STAR by multiplex PCR. A. Multiplex-tailed PCR for target region amplification. Design the primers for the left and right sides in the same arrangement for all target alleles. There is a common tail sequence on the 5’ side of the primers of the multiplex PCR followed by indexing-PCR (the slanted part of the primer). To avoid amplification by Left_Rv and Right_Fw primers, multiplex PCR for the left- and right-side primers is performed separately. B. Indexing-PCR for adding the index sequence by PCR.

### Indexing-PCR and sequencing by MiSeq

For indexing-PCR, barcodes for multiplex sequencing by MiSeq were added to each sample of the multiplex PCR amplicon using the Nextera XT Index Kit v2 (Illumina, Inc., CA, USA) and 2X KAPA HiFi HotStart ReadyMix (Nippon Genetics Co., Ltd., Tokyo, Japan). Indexing-PCR was conducted at 95°C for 3 min, followed by 8 cycles of 95°C for 30 s, 55°C for 30s, 72°C for 30s, and then 72°C for 5 min. After purification, the pooled library was sequenced with 350-bp/250-bp paired-end reads on the MiSeq platform using the MiSeq reagent kit v3 600 cycle kit (Illumina).

### Allele identification from the multiplex PCR amplicon sequencing data

Figure 2 shows an internal analysis pipeline that was constructed from an open-source software installed on our laboratory server machine. The template DNA-specific sequence of each primer on read was trimmed using Trimmomatic v.0.39^23^. Before SPAdes v.3.15.3^24^ for *de novo* assembly, aligned reads to each allele database were determined by BBtools (https://jgi.doe.gov/data-and-tools/software-tools/bbtools/). Subsequently, the identification allele number of the MLST, NG-MAST, and NG-STAR alleles was performed by similarity analysis using the Basic Local Alignment Search Tool (BLAST)^25^. These allele sequence databases were obtained from PubMLST (https://pubmlst.org/organisms/neisseria-spp, September 2022). Short and partial *gyrA* and *parC* sequences were obtained by mapping them to reference sequences using the BWA sw option^26^ and then determining the consensus sequences using Samtools^27^. quinolone-resistance determining region (QRDR) analysis was performed by the PointFinder tool^28^.

**Figure 2:**
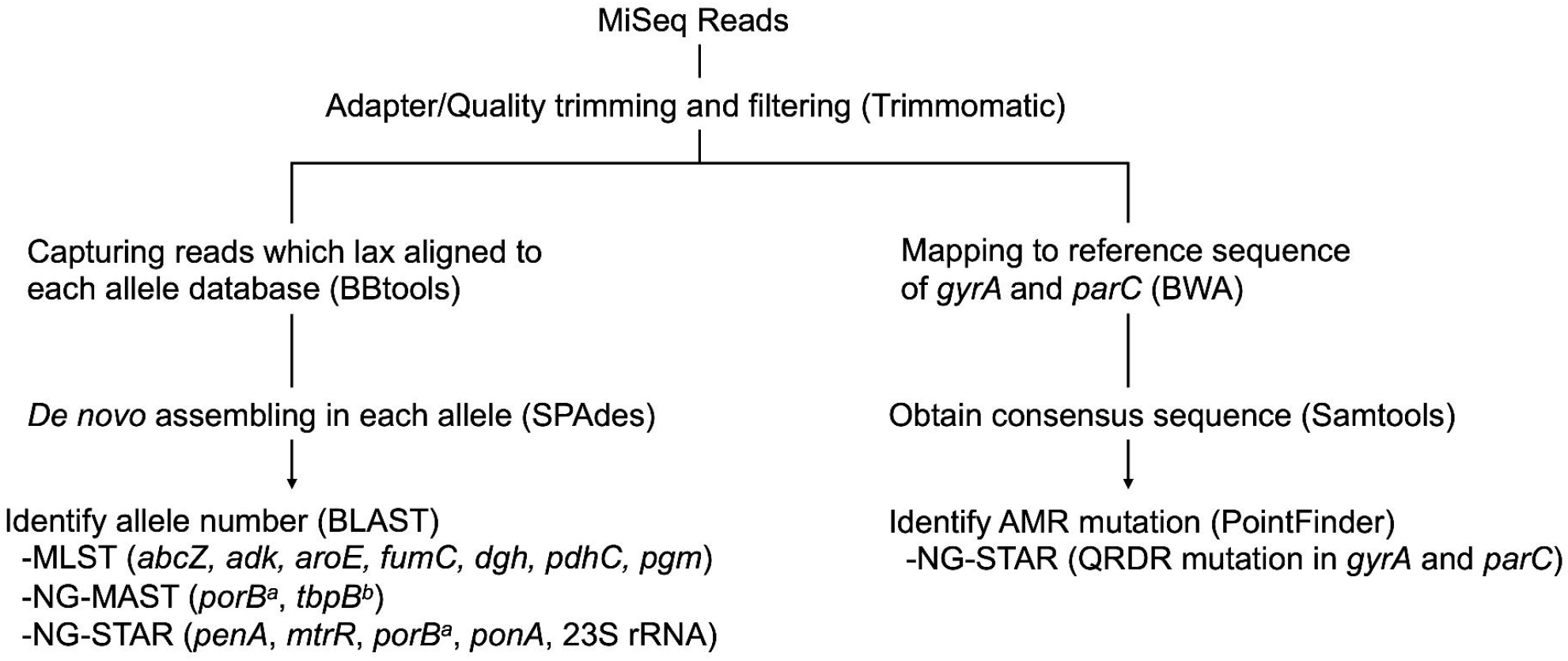
Read data analysis workflow. MiSeq read data of multiplex PCR amplicon sequencing was analyzed by a *de novo* assembly-based workflow.

### Strains for validation

To validate our novel typing method for *N. gonorrhoeae*, we used 54 of 55 strains characterized by a draft WGS (one strain was excluded because of discrepancies between the published and our reanalyzed draft WGS data)^28^. The strains contained eight MLST-ST, 38 NG-MAST-ST, 10 *penA* alleles, five *mtrR* alleles, seven *porB* alleles, two *ponA* and 23S rRNA gene alleles, and three QRDR mutation patterns detected in GyrA and ParC from translated *gyrA* and *parC* (Tables 2 and 3).

**Table 1.**
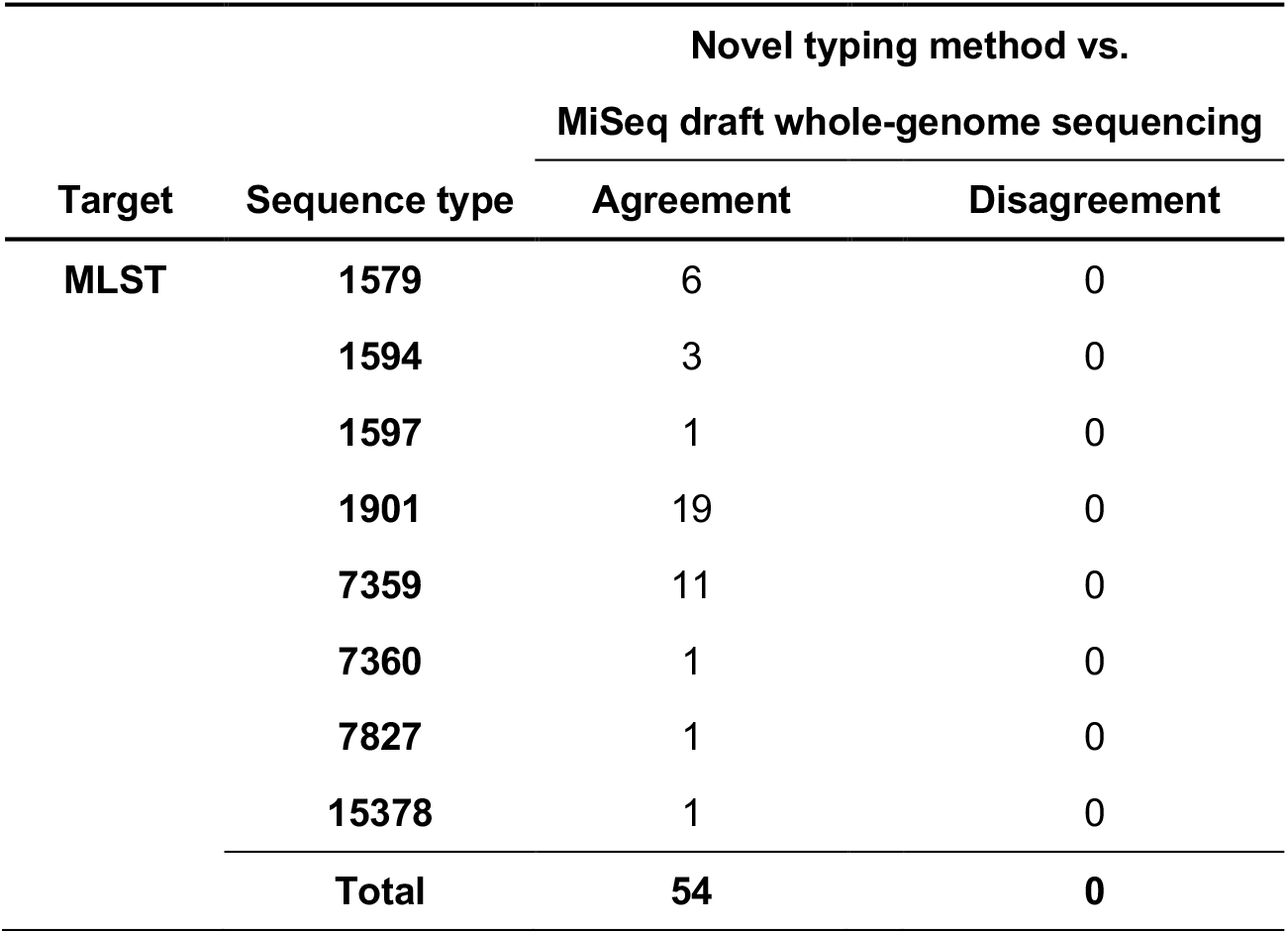
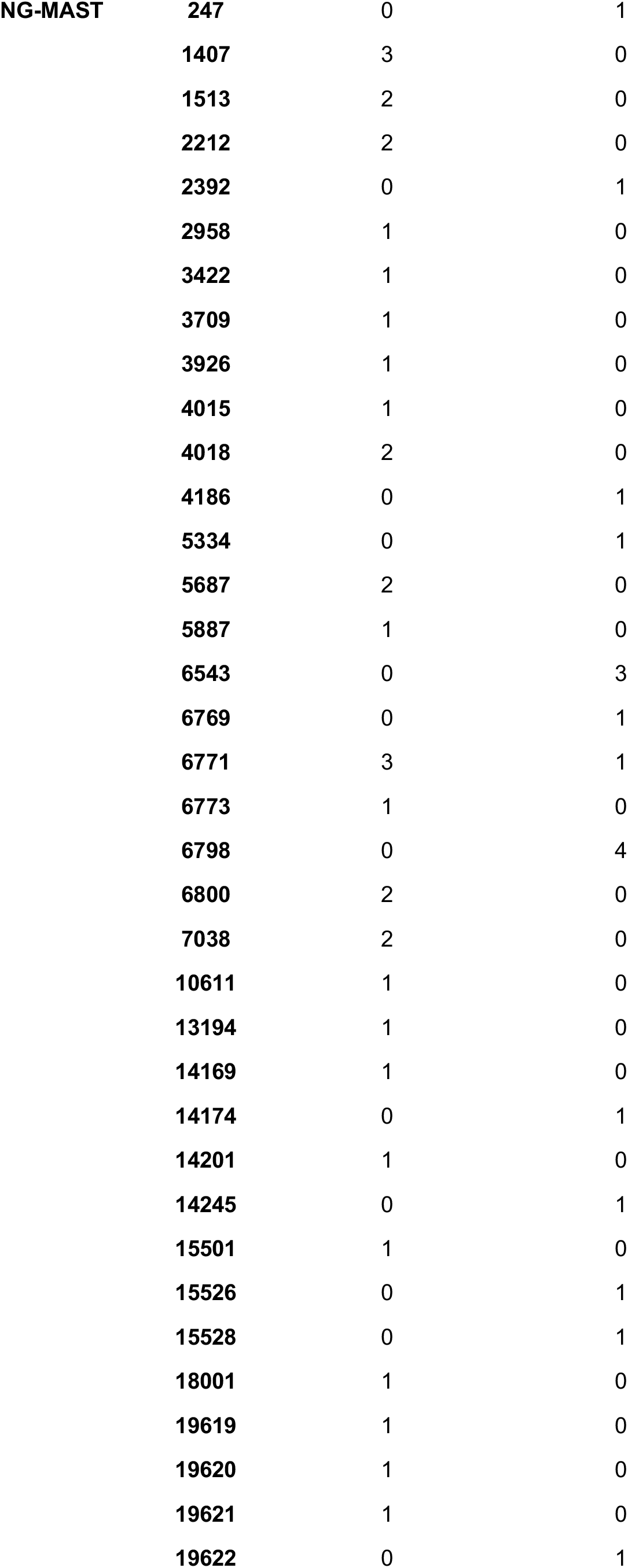
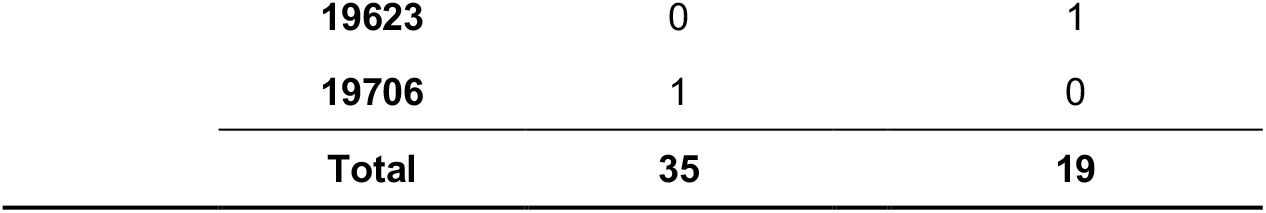
MLST and NG-MAST agreement in our novel typing method versus draft whole-genome sequencing. MLST, multilocus sequence typing; NG-MAST, *N. gonorrhoeae* multi-antigen sequence typing

**Table 2.**
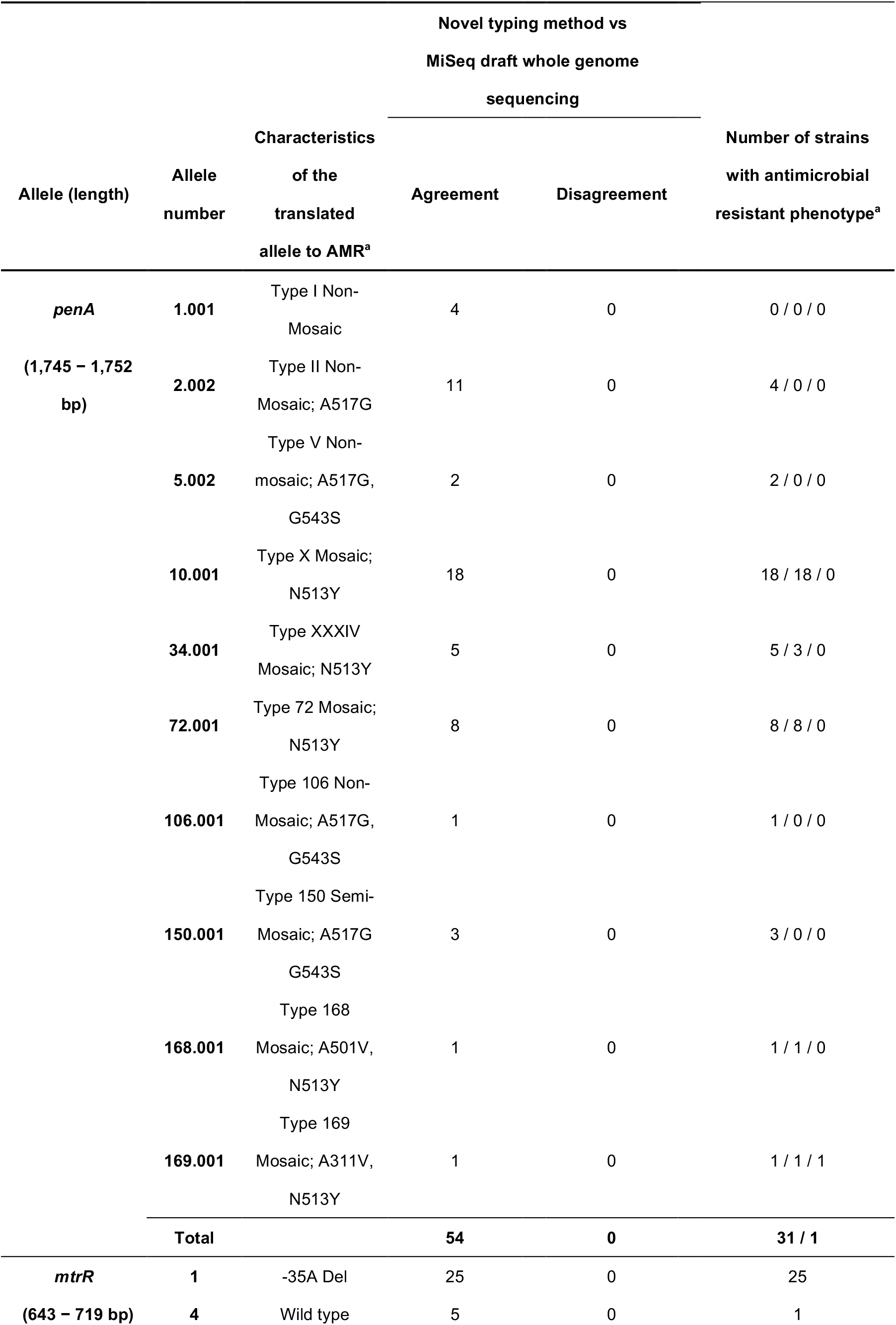
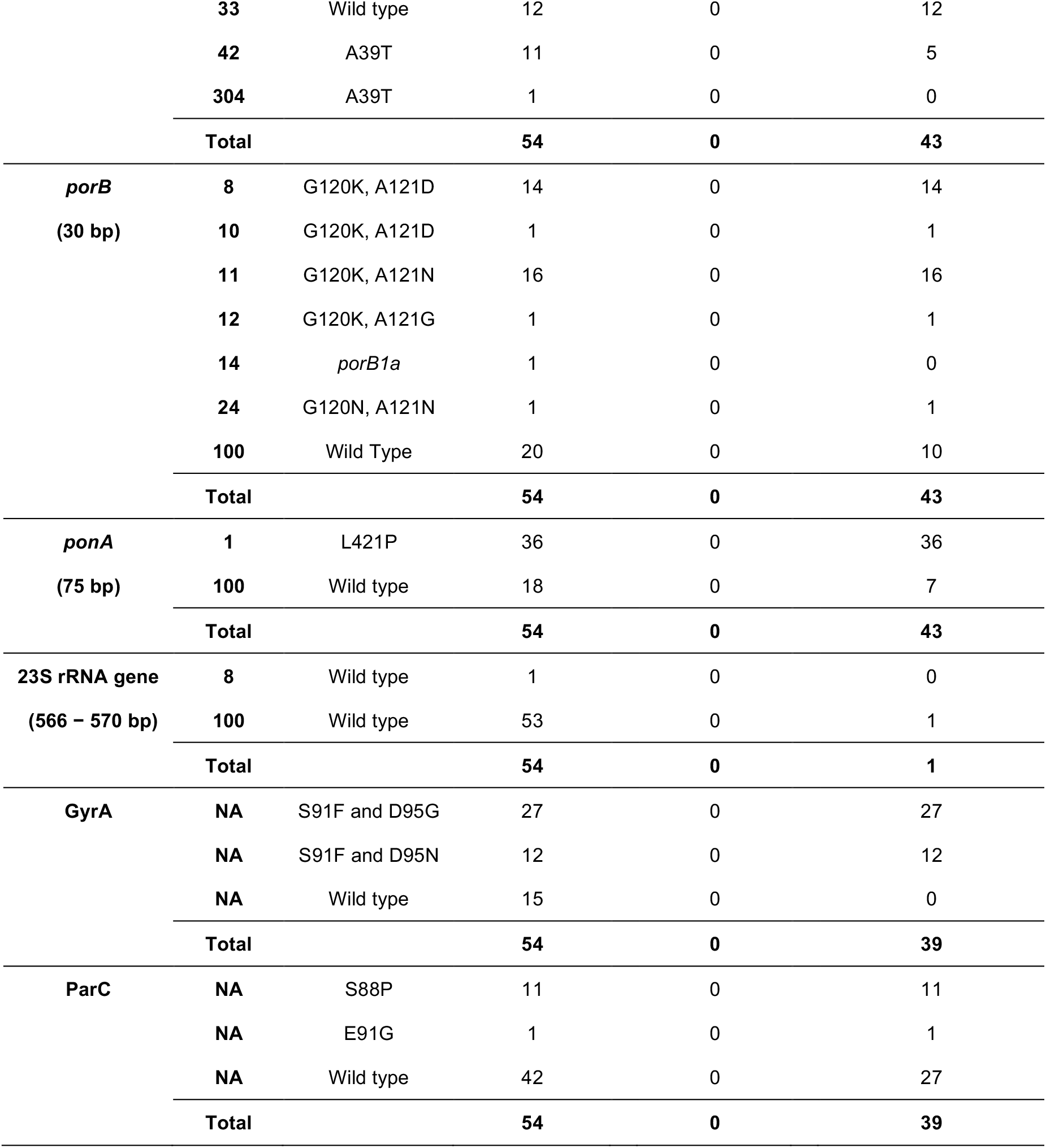
Validation of NG-STAR by our novel typing method versus draft whole-genome sequencing and antimicrobial resistant phenotype prediction. ^a^ Relationship between alleles and antimicrobial resistant phenotype: *penA*, penicillin G reduced susceptibility (>0.25 mg/L) / cefixime reduced susceptibility (>0.125 mg/L) / ceftriaxone reduced susceptibility (>0.125 mg/L); 23S rRNA, azithromycin non-susceptible (>1 mg/L); *mtrR, porB* and *ponA*, penicillin G reduced susceptibility (>0.25 mg/L); GyrA and ParC, levofloxacin resistant (>1 mg/L)^15^. NG-STAR, *N. gonorrhoeae* sequence typing for antimicrobial resistance; a, according to NG-STAR metadata; Del, deletion; amino acid abbreviations: A, alanine; D, aspartic acid; E, glutamic acid; F, phenylalanine; G, glycine; K, lysine; L, leucine; N, asparagine; P, proline; S, serine; T, threonine; V, valine; Y, tyrosine.

**Table 3.** Cause of disagreement in NG-MAST of *porB* allele with draft whole-genome sequence.

| Strain | Expected *porB* | Detected *porB* | Detection results |  |
| --- | --- | --- | --- | --- |
| ID | allele (draft | allele (top hit by | for the top hit | Causes of false allele detection |
| (TUM) | WGS) | BLAST) | allele |  |
| 15752 | 182 | 11317 | Single nucleotide mismatch | *porB* 182 is three bases shorter on the 3' end than *porB* 11317 in the NG-MAST database |
| 15755 | 4028 | 4028 | Single nucleotide mismatch | Likely due to PCR error |
| 15762 | 206 | 11889 | Single nucleotide mismatch | *porB* 206 is three bases shorter on the 3' end than *porB* 11889 in the NG-MAST database |
| 15763 | 2569 | 12073 | Single nucleotide mismatch | *porB* 2569 is three bases shorter on the 3' end than *porB* 12073 in the NG-MAST database |
| 15767 | 3854 | 11337 | Single nucleotide mismatch | *porB* 3854 is three bases shorter on the 3' end than *porB* 11337 in the NG-MAST database |
| 15770 | 4033 | 11424 | Single nucleotide mismatch | *porB* 4033 is three bases shorter on the 3' end than *porB* 11424 in the NG-MAST database |
| 15775 | 3854 | 11337 | Single nucleotide mismatch | *porB* 3854 is three bases shorter on the 3' end than *porB* 11337 in the NG-MAST database |
| 15777 | 4061 | 9083 | Single nucleotide mismatch | Likely due to PCR error |
| 15778 | 3854 | 11337 | Single nucleotide mismatch | *porB* 3854 is three bases shorter on the 3' end than *porB* 11337 in the NG-MAST database |
| 15782 | 206 | 11889 | Single nucleotide mismatch | *porB* 206 is three bases shorter on the 3' end than *porB* 11889 in the NG-MAST database |
| 15790 | 4033 | 11424 | Single nucleotide mismatch | *porB* 4033 is three bases shorter on the 3' end than *porB* 11424 in the NG-MAST database |
| 15793 | 11399 | 4069 | Single nucleotide mismatch | *porB* 11399 is six bases shorter on the 3' end than *porB* 4069 in the NG-MAST database |
| 15794 | 2547 | 11165 | Single nucleotide mismatch | *porB* 2547 is three bases shorter on the 3' end than *porB* 11165 in the NG-MAST database |
| 15796 | 4033 | 11424 | Single nucleotide mismatch | *porB* 4033 is three bases shorter on the 3' end than *porB* 11424 in the NG-MAST database |
| 15798 | 4033 | 11424 | Single nucleotide mismatch | *porB* 4033 is three bases shorter on the 3' end than *porB* 11424 in the NG-MAST database |

### Counting average sequence reads depth

The multiplex-PCR amplicon sequence reads were mapped using the BWA “sw” option to each allele sequence obtained by *de novo* assembly. Then, the sequence depth in the BAM file was determined using the pileup function in BBtools (https://jgi.doe.gov/data-and-tools/software-tools/bbtools/).

### Nucleotide sequence accession numbers

The MiSeq sequencing reads were deposited in the National Center for Biotechnology Information (NCBI) under BioProject number PRJNA901191. The Sequence Read Archive (SRA) accession number of fastq data of each sample were from SRR22270692 to SRR22270746 and are shown in Table S2.

## Results

### Evaluating sequencing read depth of each allele

The statistical range of the average sequence depth in all 15 alleles (*porB* for NG-MAST and NG-STAR was calculated as one) of each strain was as follows: mean = 425x-28,423x; median = 348x-28,320x; and standard deviation = 285x-17,901x. Sequence depth plots of each allele are shown in Figure 3.

**Figure 3.**
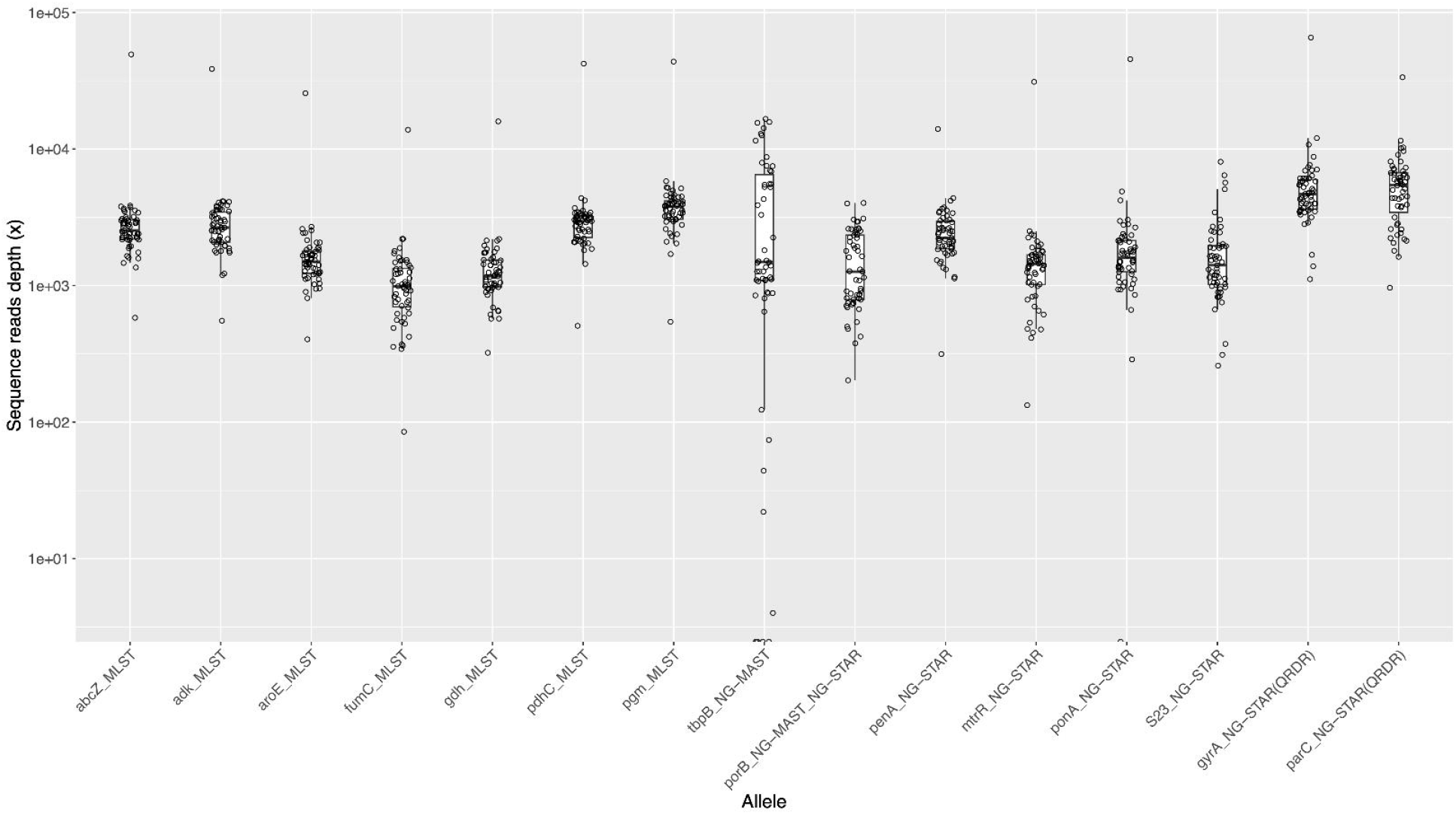
Sequence depth on each allele. Sequence depth of our novel method using MiSeq is shown as a box plot. The horizontal line in the box indicates the median, the upper end indicates the third quartile, and the lower end indicates the first quartile. The upper end of the whisker is drawn to the largest data point within 1.5× of the interquartile range (IQR) from the third quartile, while the lower end of the whisker is drawn to the smallest value within 1.5× of the IQR from the first quartile. This boxplot was generated by R ggplot2.

### Validating our method using a draft WGS as a reference

For the 54 strains of *N. gonorrhoeae* evaluated in our study, our novel typing method was in agreement with the draft WGS method using MiSeq in all alleles of MLST and NG-STAR (Tables 1 and 2). The NG-MAST results of 35 strains agreed with the draft WGS (Table 1). When evaluating the NG-MAST sequence type (ST), only one strain (NG-MAST-ST6771) both agreed and disagreed with the draft WGS data (three agreements and one disagreement) (Table 1).

### Predicting antimicrobial resistance

All strains harboring amino acid substitution patterns of S91F and D95G or S91F and D95N in GyrA were levofloxacin resistant (Table 2). Harboring a *penA*10.001 allele was associated with reduced susceptibility to cefixime but not to ceftriaxone. A strain with a *penA*169.001 allele was resistant to both cefixime and ceftriaxone. All strains harboring *penA*10.001, *penA*72.001, *penA*106.001, *penA*150.001, *penA*168.001, *penA*169.001, *mtrR*1, *mtrR*33, *porB*8, *porB*10, *porB*11, *porB*12, *porB*24, and *ponA*1 alleles showed resistance or reduced susceptibility to penicillin G. We did not observe an association between the 23S rRNA allele and azithromycin susceptibility.

### Characterizing the disagreement with the draft WGS

We identified two causes of disagreement in *porB* allele detection (Table 3). First, for 13 of 15 strains, the BLAST analysis results for detecting an exact match allele were confounded by variable allele lengths in the NG-MAST database (unlike the MLST database) leading to ambiguity. Second, we detected a single nucleotide difference at different positions in 2 of the 15 strains (TUM15755 and TUM15777).

We also determined two causes of disagreement in *tbpB* allele detection (Table 4). First, there were between two and seven mismatches in multiple *tbpB* PCR primers among five of the seven strains, which prevented the acquisition of *tbpB* contigs for these strains. Second, we found that the contig of *tbpB* was divided into two contigs in these seven strains.

**Table 4.** Primer mismatch disagreement of the *tbpB* allele between NG-MAST and the draft whole-genome sequence.

| Strain ID<br>(TUM) | Expected<br>*tbpB* allele<br>(draft WGS) | Detection<br>results | Number of mismatches on primers against<br>MiSeq draft WGS data |  |  |  |
| --- | --- | --- | --- | --- | --- | --- |
|  |  |  | left<br>(forward) | left<br>(reverse) | right<br>(forward) | right<br>(reverse) |
| 15752 | 21 | Contig divided into two | 0 | 1 | 0 | 0 |
| 15756 | 1365 | No contig generation in *tbpB* | 0 | 6 | 7 | 5 |
| 15759 | 27 | No contig generation in *tbpB* | 0 | 0 | 2 | 5 |
| 15764 | 2532 | No contig generation in *tbpB* | 5 | 6 | 5 | 5 |
| 15782 | 2528 | No contig<br>generation in<br>*tbpB* | 0 | 2 | 5 | 0 |
| 15789 | 1127 | No contig<br>generation in<br>*tbpB* | 0 | 4 | 3 | 2 |
| 15794 | 21 | Contig divided<br>into two | 0 | 1 | 0 | 0 |

## Discussion

The successful performance of MLST and NG-STAR of our developed high-throughput typing method was validated in this study. By allowing the analysis of a large number of *N. gonorrhoeae* strains concurrently, this method is both cost-effective and efficient. Furthermore, this method can potentially be used for typing *N. gonorrhoeae* and predicting AMR phenotype without culture from gonorrhea patients because of the PCR amplification specificity of each allele. Although this method is not intended to evaluate all aspects of *N. gonorrhoeae*, it will likely be a valuable tool in large-scale AMR *N. gonorrhoeae* surveillance.

Even though allelic analysis of MLST and NG-STAR was an exact match, NG-MAST did not produce the expected results with a small number of strains. There are several reasons that may explain this discrepancy. First, false higher than expected similarity scores for the allele were calculated by the BLAST analysis, which was the primary cause of the poor performance for the *porB* alleles in our study. These false higher similarity scores are likely due to the inconsistent lengths of the sequences registered in the NG-MAST database. A single nucleotide difference in a longer sequence will result in a higher similarity score than an exact match of a shorter sequence. Second, the *porB* disagreement in TUM15755 and TUM15777 is likely related to accidental nucleotide amplification errors in the multiplex-tailed PCR. The sequencing depth of *porB* was sufficient (more than 100×, Figure 3), so it is unlikely that sequencing errors were the cause. There is an option to use PCR enzyme with higher proofreading performance to prevent sequence amplification error in PCR, but at the risk that a slight primer sequence mismatch could result in a significant decrease in amplification reaction efficiency, Third, two types of poor contig formation at the *tbpB* allele may be due to primer mismatches against the allele and its surrounding sequences in the affected strains that resulted in inefficient PCR amplification. This indirectly demonstrates the low conservation of the allele sequence. This is not surprising because although NG-MAST has a higher discrimination power than MLST, it makes primer design difficult to amplify all alleles. Recently, a novel AMR *N. gonorrhoeae* was reported without NG-MAST analysis^29^, suggesting that NG-MAST may no longer be necessary in typing *N. gonorrhoeae*. We believe that WGS, not NG-MAST, is required for high resolution gonococcal transmission analysis^30^.

Because Japan has been an epicenter for AMR *N. gonorrhoeae*^12,15,31–34^, it is appropriate to use Japanese strains to validate our typing methodology. However, since the validation of the primer region mismatch is insufficient, it may be necessary to update multiplex-tailed PCR primers and that PCR conditions while using this method in the future. We are in process of applying and analyzing this method to a historical collection constructed by 220 strains isolated in Japan between 1972 and 2005. The preliminary results have been mostly favorable (manuscript in preparation).

Notably, this method of NG-STAR for *gyrA* and *parC* only targets a narrow sequence around the QRDRs and thus does not allow for allele identification of *gyrA* and *parC*. Because the goal of NG-STAR is to detect mutations that contribute to AMR, the allele number does not necessarily have to be identifiable. The idea behind this is to keep the number of PCR products as small as possible to minimize the vulnerability of the multiplex-tailed PCR. If long read sequencers become available, the vulnerability of this method would be reduced as the multiplex-tailed PCR, which is currently divided into left and right assays, could be integrated into a single assay.

In conclusion, we have developed a cost- and labor-saving high throughput *N. gonorrhoeae* genotyping method to add molecular epidemiological information to strains collected by AMR *N. gonorrhoeae* surveillance. By avoiding the sequencing of unnecessary regions in the three schemes, this method has succeeded in increasing the number of strains that can be analyzed in one MiSeq assay by approximately 5-fold compared with that obtained by WGS. This method will be valuable in AMR *N. gonorrhoeae* as it is suitable for initial analyses and can quickly identify new strains for subsequent WGS analysis.

## Data Availability

All sequencing data produced are available under the National Center for Biotechnology Information under BioProject number PRJNA901191.

## Acknowledgment

This research was supported in part by AMED under Grant Numbers JP15fk0108014h0001, JP18fk0108062j0001, and JP21fk0108605j0001.

## Notes

### Competing Interest Statement

The authors have declared no competing interest.

